# Neural correlates of insomnia with depression and anxiety from a neuroimaging perspective: A systematic review

**DOI:** 10.1101/2024.10.22.24315964

**Authors:** Chen Peng, Kai Wang, Jinyu Wang, Rick Wassing, Simon B. Eickhoff, Masoud Tahmasian, Ji Chen

**Author notes:** Contributed equally (Masoud Tahmasian, Ji Chen). Corresponding author: Center for Brain Health and Brain Technology, Global Institute of Future Technology & Institute of Psychology and Behavioral Science, Shanghai Jiao Tong University, Shanghai 200240, China. Email address (J. Chen). Corresponding author: Department of Neurology, The First Affiliated Hospital of Anhui Medical University, Anhui Medical University, Hefei 230032, China. Email address (K. Wang).

## Abstract

Insomnia affects a substantial proportion of the population and frequently co-occurs with mental illnesses including depression and anxiety. However, the neurobiological correlates of these disorders remain unclear. Here we review magnetic resonance imaging (MRI) studies assessing structural and functional brain associations with depressive and anxiety symptoms in insomnia disorder (ID; n=38), insomnia symptoms in depressive and anxiety disorders (n=14), and these symptoms in the general populations (n=2). The studies on insomnia disorder consistently showed overlapping (salience network: insula and anterior cingulate cortex) and differential MRI correlation patterns between depressive (thalamus, orbitofrontal cortex and its associated functional connectivity) and anxiety (functional connectivity associated with default mode network) symptoms. The insula was also consistently identified as indicating the severity of insomnia symptoms in depressive disorder. In contrast, findings for other regions related to insomnia symptoms in both depressive and anxiety disorders were generally inconsistent across studies, partly due to variations in methods and patient cohorts. In the general population, brain regions in the default mode network provided a functional link between insomnia and depressive symptoms. These findings underscore both the shared and distinct neural correlates among depression, anxiety, and insomnia, providing potential avenues for the clinical management of these conditions.

## 1 Introduction

Insomnia is highly prevalent in the general population and is characterized by symptoms of difficultly falling and staying asleep and waking up early with daytime dysfunction. These symptoms undermine the quality and quantity of sleep and negatively impact cognition, mood, social interactions, and quality of life[1]. Symptoms of insomnia experienced ≥3 times per week and persisting for 3 consecutive months with significant disruption to daytime activities is diagnosed as chronic insomnia disorder (ID). The current diagnostic criteria no longer distinguish between primary and secondary insomnia; this paradigm shift acknowledges insomnia as an independent disorder that requires targeted clinical management. However, insomnia does not typically occur in isolation as it involves not only the neural circuitry governing the sleep–wake cycle but also that related to emotion (reactivity/regulation) and cognition (memory/executive function). In fact, 40% of patients with ID have comorbidities[2] such as depressive disorder, anxiety disorder, bipolar disorder, post-traumatic stress disorder, and psychotic disorders and 60% of patients with these disorders report insomnia symptoms at first diagnosis[3]. Depression and anxiety are the 2 most prevalent conditions associated with insomnia symptoms[1], which are reported by 80%–90% of patients with depressive disorder[4,5] and nearly two-thirds of those with anxiety disorder[6].

The co-occurrence of insomnia symptoms with depressive or anxiety symptoms has a genetic basis, as previous genome-wide association studies in these patient populations have identified multiple overlapping genetic loci[7,8]. At the molecular level, the overlap between insomnia, depression, and anxiety may be explained by altered neurotransmission of gamma-amino butyric acid (GABA) and changed availability and expression of brain-derived neurotrophic factor. Thus, the overlapping clinical manifestations of insomnia and depressive or anxiety disorder would be expected, due to potentially shared pathophysiologic mechanisms[9]. Importantly, patients with depressive or anxiety disorder who develop persistent insomnia have poor prognosis[1,10]. Conversely, insomnia may be a prodromal symptom of depressive and anxiety disorders and has been shown to increase the risk of these disorders[11,12]; sleep medications and cognitive behavioral therapy for insomnia are commonly administered to patients and can diminish the severity of depressive and anxiety symptoms[13,14]. Additionally, good sleep hygiene can reduce the severity and residual symptoms of depression and anxiety and lower the risks of a first depressive episode and treatment-resistant depression[15].

The above evidence suggests a common neurobiological basis for insomnia, depression, and anxiety, which may be detectable in vivo by magnetic resonance imaging (MRI). Frequent cortical arousal and awakenings as well as maladaptation of amygdala reactivity—mainly during rapid eye movement (REM) sleep—are observed in insomnia disorder due to heightened activation or insufficient inhibition of the noradrenergic system, which is critical for the processing and consolidation of emotional memories[16]. Disruption of the latter process during sleep has been linked to the pathophysiologies of depression and anxiety, despite some distinct emotional states characterizing these disorders (e.g., anhedonia and fear, respectively). Indeed, MRI studies and the proposed circuit models for depression and anxiety disorder have revealed differences in their associated neurobiological patterns. For example, although both involve the medial prefrontal cortex (PFC), dopaminergic neurons projecting from the ventral tegmental area to the nucleus accumbens (NAc) modulate anhedonia[17,18] whereas fear reflects aberrant connectivity between the amygdala and ventral hippocampus[19]. Therefore, clarifying the neurobiological substrates that are common and unique to insomnia, depression, and anxiety can help guide clinical management of these disorders. However, there is a lack of overarching knowledge in this regard and there are divergent patterns of individual papers in this direction. Hence, synthesizing the current literature via a systematic review is required, as for depicting a more comprehensive picture.

Although there have been reviews of studies examining the links between insomnia and depressive or anxiety disorder, a detailed understanding of the neural correlates of these relationships based on neuroimaging evidence is still lacking. To address this gap in knowledge, this review summarized the current evidence from structural and functional MRI studies of depressive, anxiety, and insomnia symptoms and disorders. Specifically, we synthesized information on 1) depressive and anxiety symptoms in insomnia disorder, 2) insomnia symptoms in depressive and anxiety disorders; and 3) insomnia with depressive or anxiety symptoms in the general population. Besides univariate analyses based on inter-group comparisons or within-group correlations, studies that have used multivariate machine learning approaches are also included. In addition, several limitations associated with the reviewed works are indicated and a perspective for future research is provided.

## 2 Methods

### 2.1 Search strategy and study selection

This systematic review was pre-registered in the PROSPERO database (CRD42023375346) and was carried out according to Preferred Reporting Items for Systematic Reviews and Meta-analyses (PRISMA) guidelines[20]. Literature searches were conducted using Web of Science and PubMed databases. Two authors (C.P. and J.Y.W.) searched for MRI-based neuroimaging studies on insomnia, depression, and anxiety symptoms and disorders, with discrepancies independently validated by 2 other authors (J.C. and K.W.). The publication period spanned from 1980 to July 2024. Search terms in the abstract, title, or keywords were (‘depress*’ OR ‘major depressive disorder’ OR ‘MDD’) OR (‘anxiety’ OR ‘generalized anxiety disorder’ OR ‘GAD’) AND (‘Insomnia’ OR ‘Primary Insomnia’) AND ((‘magnetic resonance imaging’ OR ‘MRI’) OR (‘functional magnetic resonance imaging’ OR ‘fMRI’) OR (‘Voxel-based morphometry’ OR ‘VBM’) OR (‘structural MRI’ OR ‘sMRI’) OR (‘diffusion tensor imaging’ OR ‘DTI’)).

The retrieval process started with an automated library tool using a filter for English-language articles. MRI studies typically reported correlations between neuroimaging findings and clinical symptoms for a single disorder. The articles were grouped into 3 general categories: 1) depressive or anxiety symptoms in insomnia disorder; 2) insomnia symptoms in depression or anxiety disorder; 3) insomnia symptomatology with depressive or anxiety symptoms in the general population. Each disorder had to have been diagnosed based on official criteria (i.e., Diagnostic and Statistical Manual of Mental Disorders [DSM], International Classification of Diseases [ICD], or International Classification of Sleep Disorders [ICSD]). We excluded studies that did not report original data (e.g., case reports, reviews, meta-analyses, clinical trials) or were irrelevant to the scope of this review (e.g., those that reported on subjective sleep quality, sleepiness, or sleep disturbances other than insomnia). Additional exclusion criteria were articles focused on primary disease without neuroimaging data and neuroimaging studies of patients with mental disorders or diseases other than insomnia, depression and anxiety.

### 2.2 Data extraction and Qualitative assessment

The Participants, Inventions, Comparators, Outcomes, and Study Design (PICOS) framework[21] was used to extract the methodologic information from the selected studies including authors and year of publication, sample characteristics (size, medication, phase of the disorder, etc.), diagnostic criteria, neuroimaging modality, clinical scales, and statistical methods. Notably, as different studies have employed different versions of the diagnostic criteria with some (e.g., DSM-III-R, DSM-IV, and ICSD-2) distinguished between primary and secondary insomnia while others using the more recent versions (DSM-5 and ICSD-3) omitted these terms because of ambiguity in establishing causality between insomnia and other disorders[22], we did not make this distinction (i.e., ‘primary’ *versus* ‘secondary’) and instead adopted the terms “insomnia symptom” or “insomnia disorder” uniformly throughout the text. For clinical scales, besides the Insomnia Severity Index and Athens Insomnia Scale, we also included results based on items 4 (inability to fall asleep), 5 (waking during the night), and 6 (waking too early) of the Hamilton Depression Rating Scale insomnia subscale as a measure of insomnia severity in depressive disorders[23]. Scales and inventories used for the assessment of depressive and anxiety symptoms are shown in Supplemental Table S1, which also lists all scales/items that were used by studies included in the current review. A qualitative assessment of the included studies was conducted using a 10-point checklist to ensure the comprehensiveness of the information in the selected studies[24].

## 3 Results

### 3.1 Included studies

Selection based on the aforementioned criteria yielded 54 eligible articles, as detailed in the PRISMA flowchart (see figure 1). There were 38 studies on ID (include 2 on major depressive disorder [MDD] comorbid with ID and 2 on generalized anxiety disorder [GAD] comorbid with ID), 13 on MDD, 1 on anxiety disorder, and 2 on non-clinical, general populations. Diagnoses were based on standardized criteria; the scales used to assess symptoms along with the available information on treatments and phase of the disorder in each study are shown in Supplemental Table S1. The majority of studies (n=42) provided the information of medication whereas fewer reported the phase of the disorder (all of these were studies on MDD).

**Figure 1.**
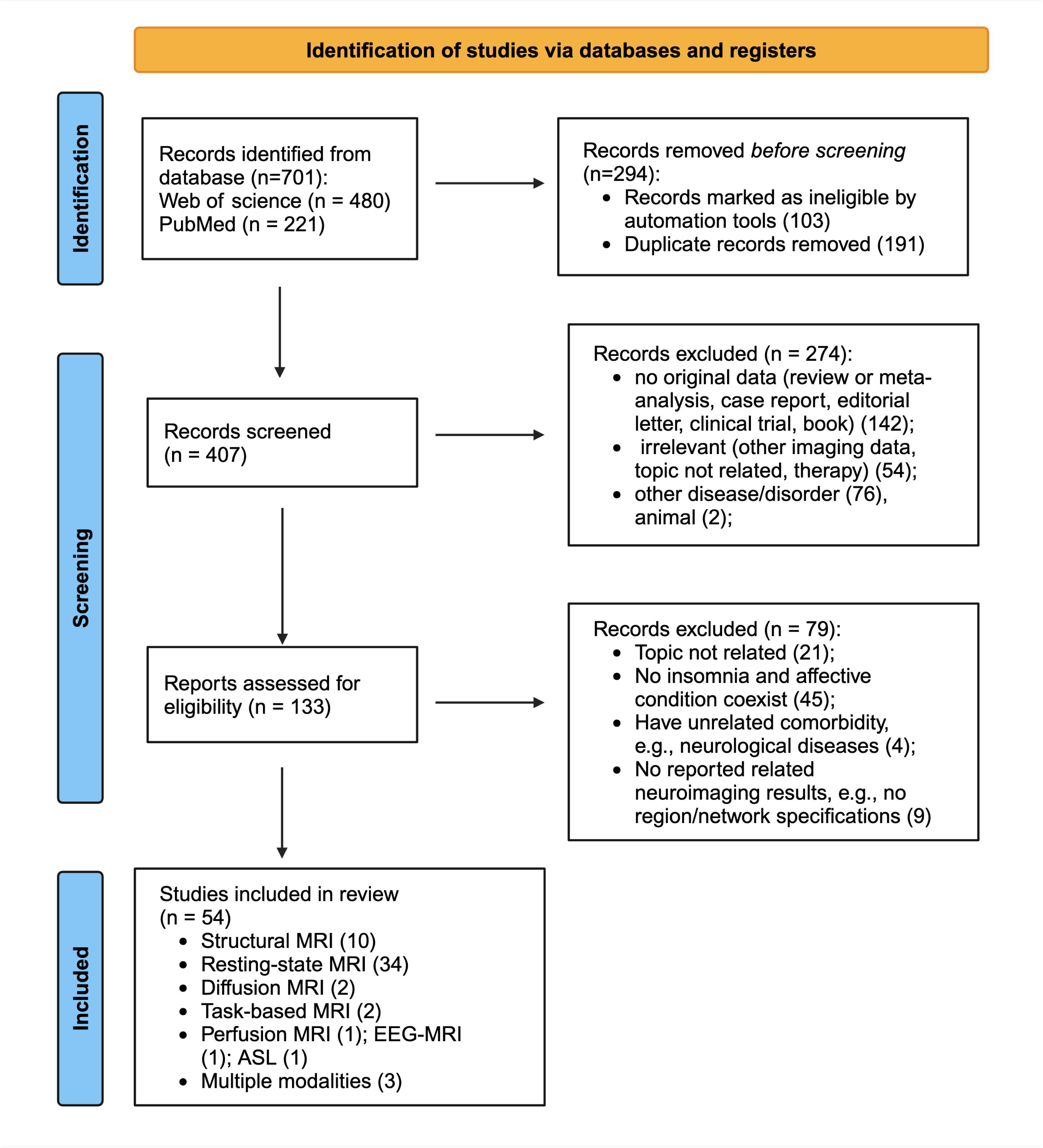
PRISMA study selection strategy **Abbreviations:** MRI: magnetic resonance imaging; EEG-MRI: simultaneous electroencephalography and MRI; ASL: arterial spin labeling MRI.

Of the included brain MRI studies, 12 provided structural metrics including grey matter morphology (10) and white matter integration (2); 34 were functional assessments (2 also reported structural measurements) and examined connectivity and spontaneous activity (regional homogeneity [ReHo] and amplitude of low-frequency fluctuations). Two studies used task-based functional MRI (fMRI) to investigate neural activation patterns during behavioral paradigms; and 3 implemented perfusion and arterial spin labeling MRI or magnetic resonance spectroscopy. Thirty-one of the 54 studies just used correlation or regression analysis to evaluate the association between neuroimaging findings and clinical scale scores for insomnia, depression, or anxiety; twelve studies involved group-level comparison approach by stratifying patients according to the level of insomnia, depression, or anxiety symptoms and compared MRI parameters across subgroups. There were 9 studies that used multivariate machine learning to identify neurological features based on prediction or classification analyses. Two studies used a mediation analysis approach.

Here we summarize the following findings for the 3 categories and their sub-conditions (if any): 1) depressive and anxiety symptoms in ID; 2) insomnia symptoms in depressive and anxiety disorder; and 3) insomnia with depressive or anxiety symptoms in the general populations. The frequency with which specific brain regions were noted as being significant across neuroimaging modalities was quantitatively analyzed (See table 2, figure 2). We defined brain regions reported ≥3 times as consistent findings whereas those reported twice required closer scrutiny, especially if the data were collected using different imaging modalities. In these cases, the overlapping and unique neural correlates across conditions were pooled.

**Figure 2.**
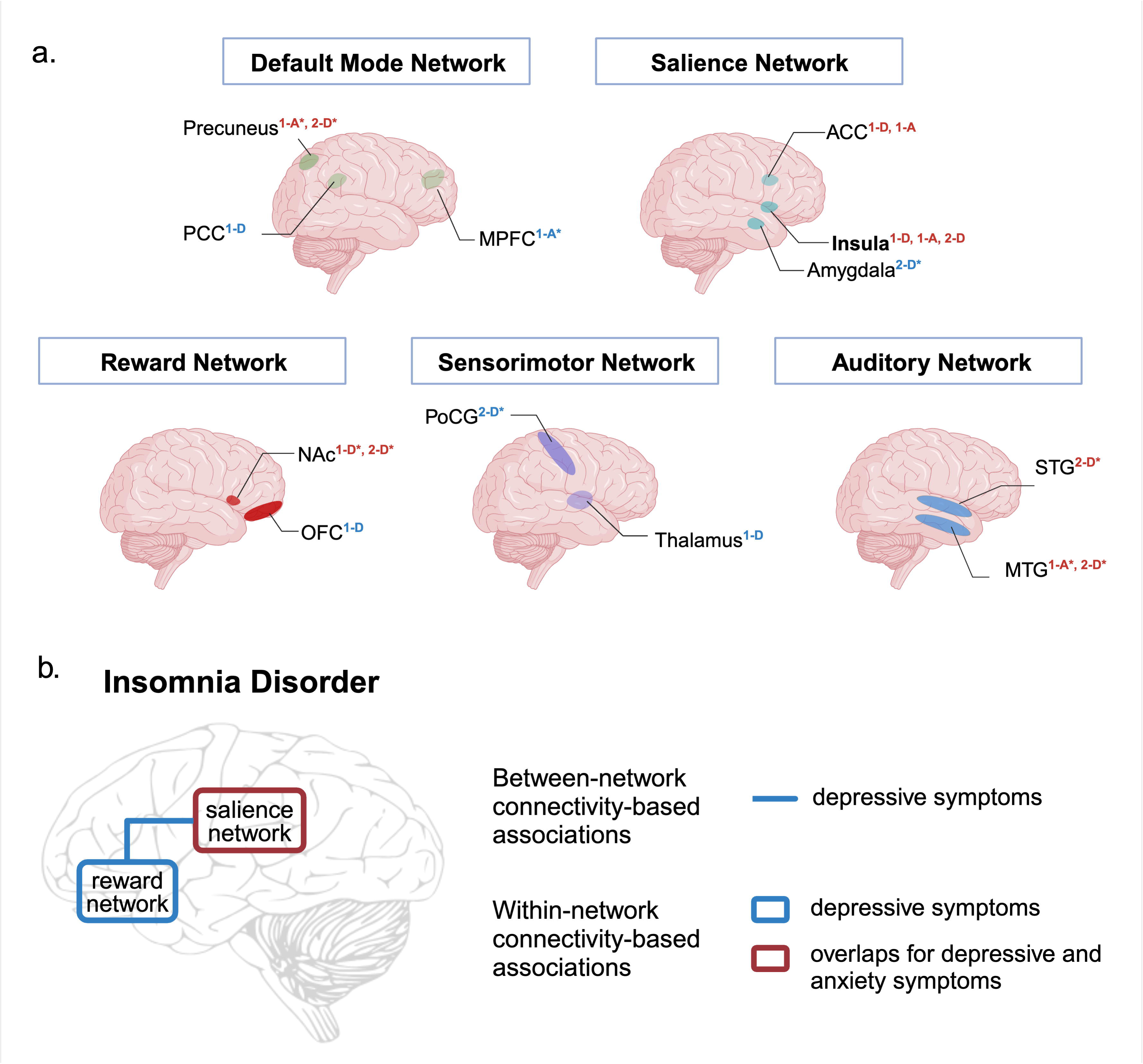
The synthesized consistent regional and connectivity-based associates of symptoms in different conditions. **a.** The synthesized consistent regional brain associates in different conditions (number of reports in >=2 studies, Created in BioRender.com.). Superscripts indicate regions having MRI metrics associated with 1: depressive (1-D) or anxiety (1-A) symptoms in ID; 2: insomnia symptoms in depressive (2-D) or anxiety (2-A) disorder. The superscripts for regions consistently reported in more than one condition are highlighted in red. Asterisk (*): regions reported twice are also indicated, for completeness and to draw attention for future works if a consistency would be expected. **b.** Synthetized model on the association of functional connectivity within-/between-brain networks consistently with depressive and/or anxiety symptoms in insomnia disorder across studies (Created in BioRender.com). Note: The red frame highlighted the overlaps regarding connectivity-based association results for depressive and anxiety symptoms in insomnia disorder; though note that the connectivity within the salience network has been reported twice in the correlation with anxiety symptoms (depressive symptoms: 3 studies). For depressive symptoms, consistent findings include also the functional connectivity within the reward network (4 studies) and the connectivity of the reward network with the salience network (3 studies). **Abbreviations:** PCC: Posterior Cingulate Cortex; MPFC: Medial prefrontal cortex; ACC: Anterior Cingulate Cortex; NAc: Nucleus accumbens; OFC: Orbitofrontal cortex; PoCG: Postcentral gyrus.

### 3.2 Neural correlates of depressive and anxiety symptoms in ID

#### 3.2.1 Depressive symptoms in ID

The severity of depressive symptoms was found to be associated with gray matter volume in the rostral anterior cingulate cortex (ACC) [25], white matter microstructural integrity in the right body of the corpus callosum[26], morphometric similarity between the paracentral lobule and the rest of the brain[27], and nodal efficiency of the left thalamus within the anatomical networks formed by white matter tracts[28].

Task-based fMRI studies revealed that activation of reward-related regions (e.g., orbitofrontal cortex [OFC] and ventral striatum [VS]) was negatively associated with depressive symptoms when subjects were presented with pictures of human faces expressing happiness/positive emotions[29]. The strength of functional connectivity in the right caudate nucleus[30] and the functional connectivity between 1) the thalamus and ACC[31]; 2) the NAc and OFC[32]; 3) the locus coeruleus (LC) and middle temporal gyrus (MTG) [33]; 4) the OFC and insula, paracingulate gyrus, and inferior parietal cortex as well as supramarginal gyrus and precuneus[34,35]; and 5) bilateral ACC[36] were shown to be correlated with the severity of depressive symptoms in ID.

Some studies examined the impact of depressive severity on insomnia disorder by stratifying the patients according to their degree of depression and comparing functional connectivity across the subgroups. The results revealed the involvement of connectivity between the NAc and the broader brain networks including the default mode network (DMN; posterior cingulate cortex [PCC]), the reward network (putamen), the salience network (insula and middle cingulate cortex [MCC]), the executive control network (dorsolateral PFC [DLPFC]) [37]; together with the sensorimotor cortex (thalamus) and visual cortex (medial occipital gyrus [MOG]). There was also reduced or increased connectivity between the ACC and left PCC or midbrain[38]. Multivariate classification analysis revealed functional connectivity between the right anterior OFC seed region and medial OFC, insula, MCC, PCC, and MTG as an important feature for distinguishing between patients with insomnia with high vs. low levels of depression[39]. Yu and colleagues (2018) [40] corroborated these findings by demonstrating the effect of an interaction between depression and insomnia on OFC gray matter volume in patients with ID with a high degree of depressive symptoms (ID-HD) and those with MDD and high degree of insomnia symptoms. The authors further found that patients with MDD and those with ID-HD had reduced cortical thickness in the bilateral superior parietal lobule, right MCC, and parahippocampal gyrus (PHG) compared with patients with ID with a low degree of depressive symptoms. Compared with healthy individuals and those with ID or MDD alone, individuals with ID and comorbid MDD had differential functional connectivity between left and right OFC, thalamus and temporal pole[41]; in these patients, decreases in cerebral blood perfusion in the cerebellum (vermis) and in functional connectivity between the hippocampus and inferior frontal gyrus (IFG) were correlated with both insomnia and depressive symptoms[42].

Despite some inconsistencies, brain MRI findings from different studies supported the important role of the OFC (6/18 studies), cingulate cortex (ACC, 4/18; MCC, 3/18; and PCC, 3/18), and the thalamus (5/18) in the manifestation of symptoms of depressive symptoms in patients with ID. Other brain regions such as the insula (2/18) and NAc (2/18) have also been reported as being relevant. The OFC, ACC, MCC, and thalamus showed both functional and structural correlates with depressive symptoms whereas the remaining studies found only functional correlates. These brain regions encompass multiple networks including the DMN and reward, salience, and sensorimotor networks. At the network level, the connectivity of the reward network especially surrounding the OFC (3/18; OFC-NAc, anterior OFC-medial OFC, left OFC-right OFC), as well as the connectivity between the reward and salience networks (3/18; including 2 studies on the connectivity between the OFC and insula) and DMN (OFC to PCC and NAc) has been reported.

#### 3.2.2 Anxiety symptoms in ID

MRI parameters in specific brain regions were associated with anxiety symptoms in patients with ID. In particular, the small basolateral nucleus of the amygdala was linked to anxiety level in these patients[43]. Other studies have described functional brain MRI correlates such as spontaneous neural activity in the ACC[44], right putamen[45], and right postcentral gyrus (PoCG)/inferior parietal lobule[46] as being significant. The global functional connectivity density (gFCD) of the left ACC and right insula was shown to be related to both insomnia and anxiety symptoms in patients with ID[47].

Functional connectivity between the ACC and precuneus[48], right PHG and left DLPFC[49], midbrain and basal forebrain[50] was positively correlated with anxiety symptoms. Additionally, stronger functional connectivity in the right dorsomedial PFC (DMPFC) was observed in patients with ID with more severe anxiety and was predictive of anxiety level[51]. Some perceptual brain regions related to elevated arousal in ID were also found to be associated with anxiety severity, especially the functional connectivity between bilateral middle occipital gyrus/posterior MTG[52] as well as the connectivity of the lateral occipital complex (LOC) and dorsal ACC with the superior fronto-parietal (central executive) network and LC seed regions, respectively[53,54]. Patients with ID presenting with anxiety symptoms showed lower long-range FCD in the precuneus and reduced functional connectivity strength between anterior (e.g., mPFC) and posterior (e.g., precuneus) DMN components compared with patients with ID and minimal anxiety[55]. In contrast to healthy control subjects, patients with ID and comorbid anxiety showed a widespread reduction in gray matter volume; the reduction in the left insula and the functional connectivity of the left cerebellum with the left MCC and medial SFG were indicative of insomnia and anxiety symptoms in these patients[56]. Another study identified a hyperarousal brain state characterized by extensive hyper-dynamic functional connectivity within and between networks. Patients with ID with comorbid GAD showed an increase in the integrated nodal strength of the PCC, and the increases in segregation and fraction of the hyperarousal state were associated with the severity of anxiety and insomnia symptoms[57].

In summary, functional brain correlates of the anxiety symptoms in patients with ID in the ACC (salience network, 4/15 studies) were the most consistent across studies, followed by the two default mode network regions: precuneus (2/15) and mPFC (2/15). Their functional connectivities (e.g., between ACC and the precuneus, ACC and LC as well as precuneus and mPFC) were commonly involved.

#### 3.2.3 Similarities and differences

According to the above, the ACC was the only brain region consistently implicated in both depressive and anxiety symptoms across studies. Some studies on ID reported additional functional connectivity for both depressive and anxiety symptoms, specifically: 1) within the salience network (insula and ACC), although the connectivity of the insula with the right fusiform nucleus and bilateral thalamus was only applicable to depressive symptoms, whereas the insula’s connectivity with the left MTG was associated with anxiety symptoms[58]; 2) between the right insula and left supramarginal gyrus[59]; and 3) between the ascending arousal network (LC, ventral tegmental area) and central executive network (posterior parietal cortex, LPFC) as well as the intraparietal sulcus and LOC[60]. Regional findings included the synchrony of ReHo in the left insula and right precentral gyrus[61], and gray matter volume reduction in the MCC[62].

Collectively, the ACC (depressive, 5/23 studies; anxiety, 5/20) and insula (depressive, 5/23; anxiety, 5/20) were consistently associated with both depressive and anxiety symptoms in ID. These brain regions are distributed in the salience network and their functional connectivity (especially that of the insula) was also shown to be related to both symptoms (salience network: depressive 9/23 studies and anxiety, 6/20; insula: depressive, 5/23, connectivity with NAc, OFC, thalamus, ACC, and the supramarginal gyrus; anxiety, 3/20, connectivity with the ACC and supramarginal gyrus as well as the gFCD of the insula). The LC (depressive 2/23, LC-MTG and LPFC; anxiety, 2/20, LC-dorsal ACC and LPFC) is a central regulatory node of the ascending arousal network that is related to the pathophysiology of insomnia; it has been less frequently reported as being involved in symptom severity of depressive and anxiety in ID, although these studies assessed functional connectivity with the LC as the seed region.

The distinct connectivity patterns for depressive and anxiety symptoms indicated the following: 1) depressive symptoms mainly involved the thalamus (5/23 studies) and the reward network (particularly the OFC [6/23] and functionally connected brain regions [4/23; NAc, insula and mOFC], and left and right OFC), and was frequently associated with the MCC (depressive, 4/23; anxiety, 2/20) and PCC (depressive, 3/23; anxiety, 1/20); and 2) anxiety symptoms were more closely linked to connectivity of the DMN (e.g., FCS of the DMPFC and precuneus, and the connectivity of the precuneus with mPFC and ACC) in ID.

### 3.3 Neural correlates of insomnia symptoms in depressive and anxiety disorder

#### 3.3.1 Insomnia symptoms in depressive disorder

Compared with patients with MDD with less severe insomnia symptoms, those with more severe symptoms exhibited the following: 1) higher spontaneous neural activity in the left precuneus[63] and anterior insula/IFG[64]; 2) higher ReHo in the PCC/precuneus, MCC, PoCG, and inferior temporal gyrus (ITG), with the activity in the PoCG and ITG reflecting the severity of insomnia symptoms[65]; 3) lower neurometabolic level (ratio of choline-containing compounds to creatinine/phosphocreatine) in the ACC[66]; 4) higher gFCD in the left ITG and posterior PHG/hippocampal gyrus[67]; and 5) higher amygdala-based functional connectivity in the bilateral superior temporal gyrus (STG) with lower connectivity in the left supplementary motor area and bilateral PoCG[68]. The functional connectivity between the bilateral amygdala and STG[68], along with increased spontaneous neural activity in the right MTG/STG[69] and insula[70] were also found to be positively associated with insomnia severity scores in patients with MDD.

Changes in brain structure—specifically, atrophy in the insular cortex and NAc[23,71,72] and increased volume of bilateral amygdala[73]—were found to be correlated with the occurrence of insomnia symptoms in MDD. Interestingly, gray matter density and spontaneous neural activity in the basal ganglia (globus pallidus, NAc, and dorsal striatum), central executive network (IFG, MFG, inferior parietal lobule), and visual network (LOC) were found to predict insomnia symptoms in patients with MDD[74].

In summary, various brain MRI parameters were reported to be correlated with the severity of insomnia symptoms in patients with depressive disorder. In particular, the most frequently overlapping findings were localized in the insula (4/13), a region that is consistently linked to both depressive and anxiety symptoms in ID, whereas others were generally inconsistent which appeared only twice across the 13 studies in total (amygdala, NAc, precuneus, PoCG, frontal (IFG) and temporal areas). Moreover, there was high variability across patient populations (in 3/13 studies, patients had medication-naïve MDD; in 4/13, some patients were medication-naïve and others had completed a washout period; and 5/13 included on-medication patients); study methods (6/13 reported both structural and fMRI findings); and the exact brain regions that were reported.

#### 3.3.2 Insomnia symptoms in anxiety disorder

Only 1 study reporting cortical complexity (local gyrification index) examined insomnia symptoms in anxiety disorder. The indices for the insula, bilateral OFC, and left MFG were negatively associated with the severity of insomnia symptoms[75].

#### 3.3.3 Similarities and differences

As there was only 1 study on anxiety disorder, a comparison with findings on the neural correlates of insomnia symptoms in depressive disorder was not possible. However, the insula was shown to be involved in all reviewed clinical conditions as summarized above.

### 3.4 Insomnia symptomatology with depressive or anxiety symptoms in the general population

Sleep disturbance often co-occurs with negative mood states. In shift workers, changes in MRI-assessed cerebral perfusion in the right PHG and cerebellum and left inferior occipital gyrus were shown to be correlated with the severity of insomnia and depressive symptoms (Hospital Anxiety and Depression Scale) [76]. A longitudinal study found that activation of the DMPFC in response to a task reward mediated the relationship between nonrestorative insomnia symptoms (feeling poorly rested upon awakening) in early adolescence and depressive symptoms emerging at later ages[77]. Although the reported regions vary, these results suggested the potentially important role of DMN.

## 4 Discussion

This review of MRI-based studies on ID found that the neurobiological ccorelates of depressive symptoms overlapped with anxiety symptoms in the insula and ACC and in the functional connectivity within the salience network pertaining to these 2 brain regions. Additionally, distinct MRI correlation patterns with depressive and anxiety symptoms were observed. The thalamus as well as the OFC and its functional connectivity within the reward network (OFC– NAc, anterior OFC–medial OFC, left OFC–right OFC) and salience network (insula) were prominently involved in depression, whereas the DMN-based connections (FCS of the DMPFC and precuneus, and the connectivity of the precuneus with mPFC and ACC) were more relevant to the severity of anxiety symptoms. While the insula was likewise consistently identified as indicating the severity of insomnia symptoms in depressive disorder, findings for other regions related to insomnia symptoms in both depressive and anxiety disorders were generally inconsistent across studies. This reflects the high heterogeneity in methodology and patient populations and highlights the need for big data analyses and machine-learning approaches to ensure reproducibility and generalizability of the findings. Additionally, we found that in the general population, insomnia and depressive symptoms were potentially linked via the DMN.

The insula is known to modulate fear and anxiety likely through GABAergic neurotransmission[78,79]. Neuroimaging and molecular analyses of postmortem brains of patients with depressive disorder has revealed dysfunction of the mu opioid receptor—which is involved in reward processing—in the anterior insular cortex among other brain regions[80]. There have been a limited number of studies in animal models examining the role of the insula in sleep and wakefulness despite its reciprocal anatomic connections with the hypothalamus and brainstem, which are importantly involved in sleep regulation[81]. There is also a leision study supported dysfunction of the anterior insula to underlie changes in sleep-wake patterns as observed in brain disorders[82]. The insula is a central node of the salience network, which mediates the DMN and central executive network, integrating cognitive, emotional, and physiologic functions based on external–internal interaction[83]. In addition to localized abnormalities, the functional connectivity of the insula (for example, with the ACC—i.e., connectivity within the salience network) is related to both anxiety and depressive symptoms in ID, although other connected brain regions—particularly the OFC—are only implicated in depressive symptoms. The ACC overlaps with both depressive and anxiety symptoms. As a key node in the salience network, the ACC is thought to integrate information for the regulation of uncomfortable emotions[84]. Importantly, the subgenual ACC was identified in our meta-analysis as a brain region that is consistently altered in patients with ID[85] and may serve as a transcranial magnetic stimulation target in the treatment of MDD[86]. Along with the MCC, the cingulate cortex may be involved in the pathophysiology of both depression and anxiety (e.g., impaired reward processing and fear response pathways).

The OFC—specifically the medial part and connections with the insula—is more closely linked to depressive symptoms than anxiety as it is important for reward processing. Failure to obtain expected rewards can lead to emotional disturbance, including feelings of sadness and depression. A dysfunctional OFC has been commonly proposed as a pathophysiologic mechanism underlying depressive disorder[87]. In addition to projecting reward-related information to the ventral striatum, the OFC also outputs to the insula[88]. This at least partially explains why the functional connectivity between the insula and OFC is linked to depressive symptoms in ID. The thalamus may provide another link between depressive severity and ID as it is part of the arousal system that regulates the sleep–wake cycle[89]. The level of anxiety was linked to the precuneus (posterior DMN) and its functional connections (e.g., with the ACC [salience network] and mPFC [anterior DMN]). The DMN is critical for emotion processing as well as introspection and self-referential processing[83]. Imbalanced anterior–posterior DMN activity and disrupted DMN–salience network connectivity have been observed in patients with ID. The former affects the normal wake–sleep cycle and affective memory consolidation during slow-wave sleep[90], whereas the latter may be responsible for impaired external and interoceptive processing in anxiety[91], although such abnormalities have also been observed in depressive disorder[92]. Moreover, in the general population, functional deficits in DMN nodes (DMPFC, PHG) mediated the relationship between insomnia symptomatology and depressive symptoms. According to the literature, individuals with DMN dysfunction are at risk for developing depression as they would tend to prioritize negative over positive information[93].

Other less frequently reported brain regions linking insomnia with depression and anxiety were the LC (depressive and anxiety symptoms in ID) and amygdala (insomnia symptoms in depressive disorder). The LC is critical for sleep and circadian rhythm regulation[94] whereas the nocturnal adaptation of amygdala reactivity is modulated by the noradrenergic system[95], which includes the LC. In patients with ID, restless REM sleep resulted in incomplete suppression of LC activity and heightened activation or insufficient inhibition of the noradrenergic system, which negatively impacted the amygdala’s emotion processing function[16]. In a rat model of stress-induced sleep disturbance, the amygdala showed aberrant activity, leading to cortical arousal via modulation of the LC and a pattern of sleep fragmentation similar to that observed in humans with acute insomnia following exposure to stress[96]. Restless sleep can in turn disrupt the coordination between the amygdala and hippocampus, hindering the normal consolidation of emotional memories and diminishing an individual’s ability to adapt to emotional distress, thereby potentially increasing depressive symptoms and negatively affecting sleep[97]. Patients with MDD and anxiety exhibit hyperactivation of the LC and the LC noradrenalinergic neurotransmission system has been proposed as a treatment target[98]. Besides the involvement of the amygdala in insomnia symptoms in MDD, nocturnal adaptation of limbic circuits may be compromised by LC and noradrenalinergic system dysfunction especially surrounding the ACC, hindering the formation and moderation of long-term emotional memory traces and the resolution of distress[84]. Over time, this may lead to a chronic state of hyperarousal and an increased risk of developing depression and anxiety. However, the ACC has not been consistently found to correlate with insomnia symptoms in depressive disorder, suggesting that insomnia perturbs emotional response and depression contributes to insomnia via distinct mechanisms, despite the overlapping neurobiological factors such as the hypothalamic–pituitary–adrenal axis and cortisol secretion and an imbalance in the aminergic and cholinergic systems[99].

Although this review provides a comprehensive summary of the results from recent studies on the neuroimaging correlates of insomnia, depression, and anxiety, caution must be exercised when considering the findings because of certain limitations to the analyses. First, the majority of functional connectivity MRI studies used seed-based approaches that may have identified a few seed regions (e.g., LC) while excluding others, although the seed regions were presumably selected based on a priori knowledge. This bias can be overcome by large-scale whole-brain analyses and connectivity-based meta-analyses. Second, the patient populations in the neuroimaging studies had heterogeneous treatment status, making it difficult to isolate the neural correlates attributable to pathophysiology from the impact of illness duration. Third, most studies had a small sample size (<200 participants) and there was mostly no validation of the results in an independent sample. Third, the cutoff scores used to define high vs. low symptom subgroups were arbitrary (given that symptom presentation is a continuum) and also varied across studies. Fourth, most studies were cross-sectional and used a univariate analytic approach, but longitudinal studies with machine learning-based prediction algorithms are warranted to identify neuroimaging markers of ID co-occurring with depression or anxiety symptoms. Finally, ID includes various subtypes that can be further complicated by the presence of depressive or anxiety symptoms, which are also on a spectrum (e.g., MDD and anxiety disorders). As such heterogeneity would mask significant associations between insomnia and depression or anxiety, investigating specific subtypes of these disorders may yield more useful information regarding their neural correlates.

## 5 Conclusion and Outlook

This systematic review summarized the current state of knowledge on the association between insomnia, depressive, and anxiety symptoms and disorders based on neuroimaging findings. Investigations on ID reported overlapping (insula and ACC) and differential MRI-based correlates of depressive (thalamus and the OFC and its functional connectivity) and anxiety (DMN-based connectivity)symptoms. However, apart from the insula, findings for other brain regions on insomnia symptoms in depressive and anxiety disorders were generally not consistent across studies, potentially owing to the high heterogeneity in study design, patient populations, and analytic (mostly univariate) approaches. Application of artificial intelligence and machine learning strategies along with big data analysis in the future could be useful for ensuring the generalizability and reproducibility of the results for a more detailed understanding of insomnia, depression, and anxiety and their interactions from a neurobiological standpoint. Additionally, open data sharing and international collaborations such as the ENIGMA-Sleep Consortium[100] could provide more opportunities to reliably identify neural correlates of insomnia, depression, and anxiety in different cultures, clinical practice settings, and medical systems as demonstrated for other brain disorders such as Alzheimer disease. Although the current findings require validation, they nonetheless provide a comprehensive view of how insomnia can interact with depression and anxiety at the neurobiological level, thus informing the development of novel targeted interventions for the treatment of these disorders.

## Practice Points

1. The literature suggests insula abnormalities are common across depressive and anxiety symptoms in insomnia disorder, and insomnia symptoms in depressive and anxiety disorders. More research is needed on insomnia symptoms in anxiety disorder for a comprehensive comparison with depressive disorder. The insula is a crucial region for regulating negative emotions and sleep patterns;
2. Despite the extant multiple studies examining the link between insomnia symptoms and depressive disorder, significant variability in study design, preprocessing and analysis choices, stage of the disorder, and treatment status has hindered the reliable identification of brain regions beyond the insula;
3. Functional brain associations with depressive or anxiety symptoms in insomnia disorder showed multiple consistent results across studies, which revealed both overlapping and divergent patterns, highlighting the role of the salience network (anterior cingulate cortex and insula), thalamus, orbitofrontal cortex, and hubs of default mode network regions. These areas could

## Research Agenda

1. Longitudinal designs are needed to delineate the relations between long-term neural changes and symptom severity in depressive, anxiety, and insomnia disorders, while shedding light on a causal link between the emergence and co-manifestation of these conditions.
2. Given the clinical heterogeneity within insomnia disorder, investigating based on its specific subtypes may yield more helpful information regarding the neural interplay with depression and anxiety conditions.
3. Future research may consider using artificial intelligence and machine learning strategies more, alongside big data analysis, to improve the generalizability and reproducibility of findings towards precision psychiatry;
4. Open data sharing and international multi-site collaborations such as the ENIGMA-Sleep Consortium would provide more opportunities to identify neural correlates of insomnia, depression, and anxiety in different ethnicities, cultures, and medical systems.

## Data Availability

All data produced in the present work are contained in the manuscript

## Abbreviations

ACC/MCC/PCC: anterior/middle/posterior cingulate cortex
DMN: default mode network
DSM: Diagnostic and Statistical Manual of Mental Disorders
FC: functional connectivity
GABA: gamma-amino butyric acid
GAD: generalized anxiety disorder
ICD: International Classification of Diseases
ICSD: International Classification of Sleep Disorders
ID: insomnia disorder
ITG/MTG/STG: inferior/middle/superior temporal gyrus
ITG/MTG/SFG: inferior/middle/superior frontal gyrus
LC: locus coerleus
LOC: lateral occipital complex
MDD: major depressive disorder
MRI: magnetic resonance imaging
NAc: Nucleus Accumbens
OFC: orbitofrontal cortex
PFC: prefrontal cortex
PHG: parahippocampal gyrus
PoCG: postcentral gyrus
ReHo: regional homogeneity
REM: rapid eye movement
VS: ventral striatum

## Acknowledgments

This work was supported by the STI2030-Major Projects (No. 2022ZD0214000 [to JC]), the National Key R&D Program of China (No. 2021YFC2502200 [to JC]), the National Natural Science Foundation of China (No. 82371506 [to JC] & No. 82201658 [to JC]), and the Helmholtz Imaging Platform grant (NimRLS, ZT-I-PF-4-010 to SBE).

---

**Table 1.**
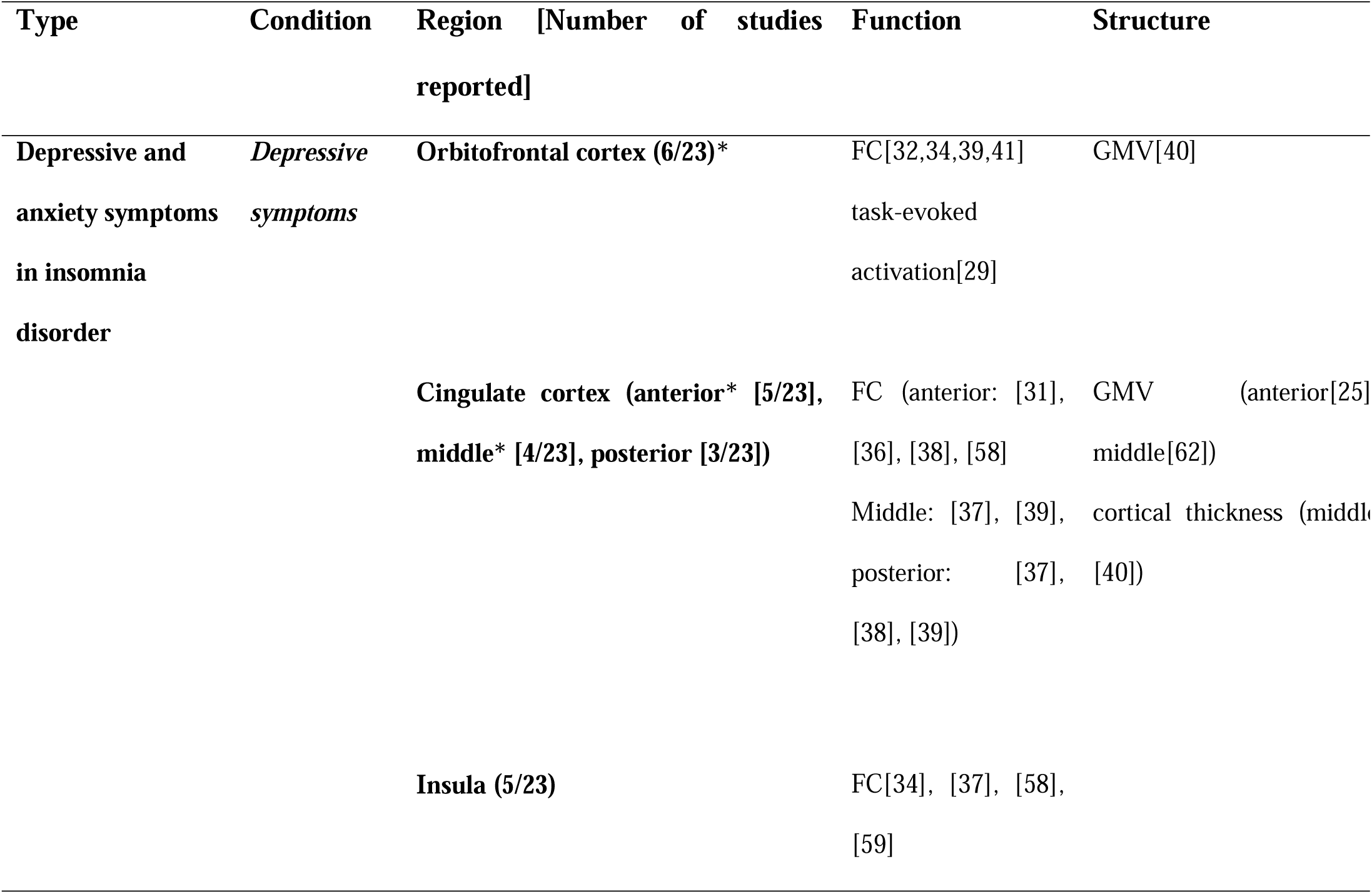
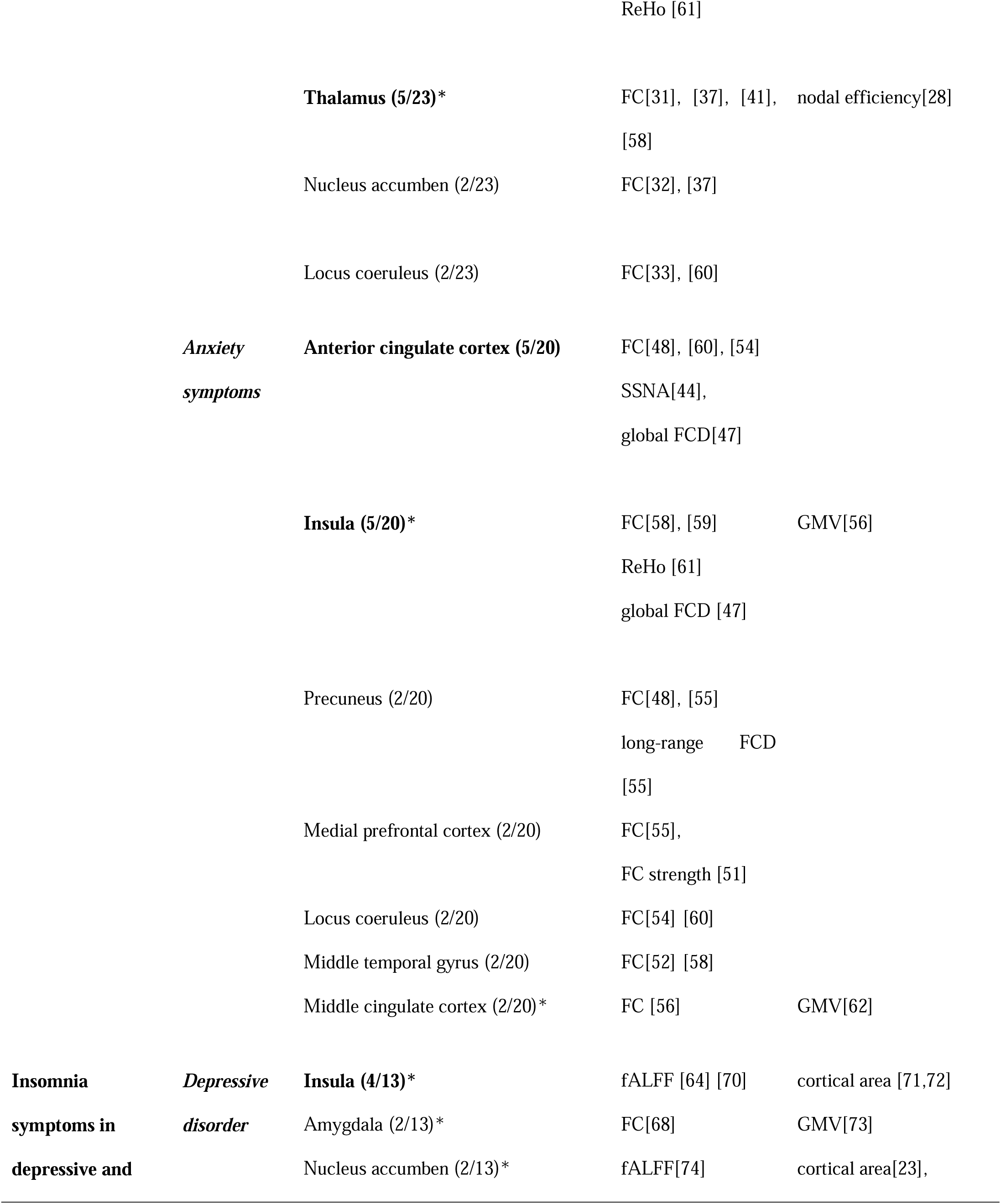
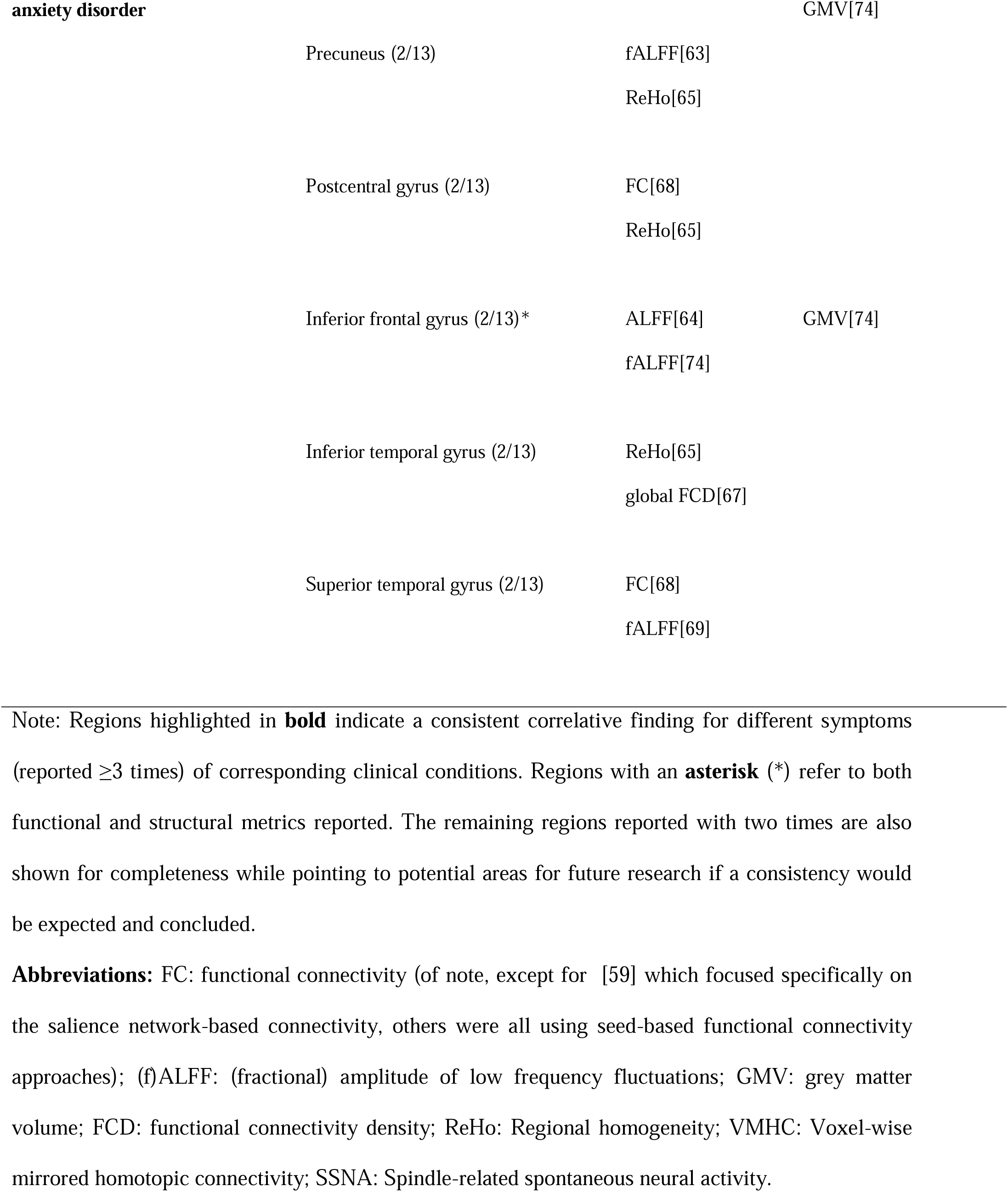
Summary of regions reported for an association with symptoms in different conditions.

