## Supplemental Table 1 for "Neural correlates of insomnia with depression and anxiety from a neuroimaging perspective: A systematic review"

**Table S1. Final studies included in the systematic review.**

Note:

ID: Insomnia Disorder, HC: Healthy Control, sMRI: structural MRI, rsfMRI: resting-state functional MRI, DTI: Diffusion Tensor Imaging;

*-HD: * with high level depressive symptoms, *-LD: * with low level depressive symptoms, *-HI: * with high level insomnia symptoms, *-LI: * with low level insomnia symptoms;

-: not reported, x: no use;

| **Study** | **Statistics (comorbidity-related)** | **Participants** | **Diagnostic Criteria** | **Medication** | **Phase** | **Scale (sleep, depression, anxiety)** | **Neuroimaging Modality** | **10-point checklist** |
| --- | --- | --- | --- | --- | --- | --- | --- | --- |
| Vassilopoulou et al., 2013[73] | Regression | 22 melancholic MDD  17 psychotic MDD  18 HC | DSM-IIIR | All for MDD | Single episode /recurrent | HAMD | sMRI | 10 |
| Ye et al., 2023[68] | Between-group comparison Correlation | 56 MDD-LI  46 MDD-HI  57 HC | DSM-V, HAMD-17>17 | Medication-free or wash out at least 4 weeks prior | - | HAMD-17 *cutoff for insomnia level:  high level: HAMD (sleep) >= 4 low level: HAMD (sleep) < 3 | rsfMRI | 10 |
| Gong et al., 2020[67] | Between-group comparison Correlation | 110 MDD  38 HC | DSM-V,  HAMD-17>=17  HC: HAMD <=7 | naïve to antidepressant medication / washout period at least five half-lives | - | HAMD-17  *cutoff for insomnia level:  high level: HAMD (sleep) >= 4 low level: HAMD (sleep) < 3 | rsfMRI | 9 |
| Yan et al., 2018[36] | Correlation | 26 ID  28 HC | DSM-IV | psychoactive medication not taken for at least 2 weeks before | - | PSQI, SAS, SDS, ISI | rsfMRI | 10 |
| Wu et al., 2018[70] | Correlation | 44 ID  46 HC | DSM-V, SDS SAS <70 | - | - | PSQI, ISI, SAS, SDS | DTI | 9.5 |
| Li et al., 2017[34] | Correlation | 50 ID  40 HC | DSM-IV | - | - | PSQI, ISI, SAS, SDS | rsfMRI | 10 |
| Li et al., 2019[31] | Correlation | 27 ID  39 HC | DSM-IV | at least 3 months no medications | - | PSQI, ISI, SAS, SDS | rsfMRI | 10 |
| Wang et al., 2023[48] | Correlation | 57 ID  46 HC | DSM-IV | Medication-naive | - | PSQI, SDS, SAS | rsfMRI | 9.5 |
| Li et al., 2018[62] | Correlation | 60 ID  53 HC | DSM-IV | free of any psychoactive medication at least 2 weeks | - | PSQI, ISI, SAS, SDS | sMRI | 9 |
| Li et al., 2018[53] | Correlation | 59 ID  53 HC | DSM-IV | free of any psychoactive medication for at least 2 weeks | - | PSQI, ISI, SAS, SDS | rsfMRI | 9.5 |
| Gong et al., 2019[43] | Correlation | 65 ID  55 HC | DSM-V, ICSD-3 | Medication-naive | / | PSQI, SDS, SAS | sMRI | 10 |
| Chu et al., 2023[71] | Correlation | 72 melancholic MDD; 74 non-melancholic MDD; 81 HC | DSM-IV, HAMD-17> 18 | Medication-naive | First-episode | Montgomery-Asberg Depression Rating Scale (MADRS); HAMD | sMRI | 10 |
| Shi et al., 2021[74] | eXtreme Gradient Boosting classification;  Correlation (ML) | Discovery set: 92 MDD  Independent;  Validation set: 460 MDD, 470 HC | DSM-4 | Discovery set: x  Validation set: - | - | HAMD | sMRI, rsfMRI | 9.5 |
| Leerssen et al., 2020[23] | Regression | ENIGMA: 1053 MDD  1277 HC  HCP: 831 HC | DSM; Schedules for Clinical Assessment in Neuropsychiatry;  International Neuropsychiatric Interview | 69% use antidepressant | First episode: 290  Recurrent: 763 | HAMD, PSQI | sMRI | 10 |
| Wu et al., 2021[70] | Regression  Correlation | 29 MDD  58 HC | DSM-IV | 25 medication- naïve, 4 medication- free (no medication for 3 months) | - | HAMD-24 | rsfMRI | 10 |
| Motomura et al., 2021[29] | Correlation | 15 ID  30 HC | DSM | 3-day washout period for hypnotics | - | PSQI, Athens Insomnia Scale (AIS), SDS | Task fMRI (Emotional-face viewing task) | 10 |
| Yang et al., 2023[35] | Correlation | 44 ID  42 HC | DSM-IV | Not use at least 2 weeks | - | PSQI, ISI, SAS, SDS | rsfMRI | 10 |
| Zhou et al., 2018[52] | Correlation | 29 ID  30 HC | DSM-V | Free of medication | - | PSQI, State Trait Anxiety Inventory-state (STAI-s), STAI-trait, BDI-II | rsfMRI | 10 |
| Jiang et al., 2022[50] | Correlation | 102 ID  96 HC | DSM-V | Not use at least 2 weeks | - | PSQI, ISI, SDS, SAS | rsfMRI | 9.5 |
| Gong et al., 2023b [37] | Between-group comparison; Multivariate linear regression  (ML) | 20 CID-HD  24 CID-LD | ICSD-3 | Not use at least 2 weeks | - | PSQI, SDS, SAS *cutoff for depressive level:  high-level: SDS>55 low-level: SDS<50 | rsfMRI | 9 |
| Li et al., 2019[57] | ICA;  Between-group comparison; Correlation  (ML) | 24 HC  15 GAD  17 GAD with insomnia symptoms | DSM-V | No medication for insomnia and anxiety | - | HAMA, STAI, HAMD-24, ISI, PSQI | rsfMRI | 9 |
| Zhou et al., 2017[46] | Correlation | 27 ID  27 HC | DSM-IV | Without medical treatment | - | PSQI, BDI-II, STAI-s, STAI-t | rsfMRI | 9.5 |
| Li et al., 2022[33] | Correlation | 49 ID  47 HC | DSM-V | Not use at least 2 weeks | - | ISI, PSQI, SAS, SDS | rsfMRI | 10 |
| Yang et al., 2022[49] | Correlation | 110 ID  60 good sleep control | ICSD-3, PSQI>7 | Not use at least 2 weeks | - | PSQI, SAS, SDS | rsfMRI | 9.5 |
| Zhang et al., 2022[75] | Correlation | 73 GAD  74 HC | DSM-V | - | - | HAMA-14, ISI | sMRI | 9.5 |
| Meng et al., 2020[45] | Correlation | 59 ID  47 HC | DSM-IV | Not use at least 2 weeks | - | ISI, PSQI, Epworth Sleepness Scale (ESS), SAS, SDS | rsfMRI | 10 |
| Wang et al., 2017[58] | Correlation | 63 ID  48 HC | DSM-IV | Not use at least 2 weeks | - | PSQI, ISI, SAS, SDS | rsfMRI | 9 |
| Winkelman et al., 2013[25] | Correlation | Study 1  ID:20 HC: 15  Study 2  ID: 21 HC: 20 | DSM-IV | Not use at least 2 weeks | - | ISI, PSQI, Dysfunctional Beliefs and Attitudes about Sleep (DBAS-16), BDI-IA | sMRI | 10 |
| Liu et al., 2018[64] | Between-group comparison; Correlation | MDD-HI: 24  MDD-LI: 37  HC: 51 | DSM-IV | 4 medication-free  57 take medication | - | HAMD, HAMA  *cutoff for insomnia level: HAMD (sleep)-3 | rsfMRI | 9 |
| Li et al., 2018[38] | Between-group comparison; Correlation | 15 ID  15 ID with depressive symptoms  15 HC | DSM-IV | Medication-naive | - | PSQI, HAMD, HAMA  *cutoff for depressive level: without: HAMD -7  With: 7<=HAMD<=24 | rsfMRI | 9 |
| Li et al., 2019[30]  Li Chao | SVM  Correlation  (ML) | 38 ID  44 HC | DSM-IV | Not use at least 2 weeks | - | ISI, PSQI, SAS, SDS | rsfMRI | 10 |
| Casement et al., 2016[77] | Mediation | 123 adolescent girls | / | - | - | The Schedule for Affective Disorders and Schizophrenia for School-Age Children (K-SADS)-insomnia&depression symptoms | Task fMRI (Reward Processing Task) | 10 |
| Xu et al., 2023[42] | Support vector regression (SVR)  Between-group comparison (ML) | 47 ID with anxiety; 49 ID without anxiety;  48 good sleeper controls (GSC) | ICSD-3 (PSQI > 7) | - | - | PSQI, ISI, SDS, SAS *cutoff for anxiety level: with anxiety: SAS > 50 without anxiety: SAS < 45 | rsfMRI | 9.5 |
| Toenders et al., 2020 [72] | K-means  (ML) | 275 MDD  100 HC | DSM-IV Axis I disorders | ~20% use antidepressant | ~30% recurrent | Montgomery-Asberg Depression Rating Scale (MADRS); Quick Inventory of Depressive Symptomatolog, Self-Report (QIDS-SR); GAD-7 | sMRI | 9.5 |
| Gong et al., 2022 [39] | Between-group comparison  SVM  (ML) | Cohort 1:  26 ID-HD,  26 ID-LD  Cohort 2:  29 ID-HD  24 ID-LD | ICSD-3 | Not use at least 2 weeks | - | PSQI, SDS, SAS *cutoff for depressive level:  high-level: SDS>55 low-level: SDS<50 | sMRI | 10 |
| Li et al., 2016[10] | linear regression | 23 ID  30 HC | DSM-IV | Not use at least 2 weeks | - | PSQI, ISI, SDS, SAS | DTI | 10 |
| Park et al., 2019[76] | Between-group comparison | 15 rotational shift workers;  15 day-workers | / | - | - | ESS, ISI, Horne-Östberg Morning-Eveningness Questionnaire, Hospital Anxiety and Depression Scale (HADS) | Perfusion MRI | 9 |
| Wang et al., 2016 [61] | Correlation | 59 ID  47 HC | DSM-IV | - | - | PSQI, ISI, SDS, SAS | rsfMRI | 10 |
| Zheng et al., 2023 [63] | Between-group comparison; Correlation | 24 MDD with sleep disturbance 33 MDD without sleep disturbance 32 HC | DSM-V | Medication-naïve (no antidepressant treatment) | - | HAMD-17, Beck anxiety inventory (BAI) | rsfMRI | 9 |
| Shao et al., 2021[44] | Correlation | 16 ID  30 HC | DSM-V, ISI>=15 | - | - | PSQI, ISI, SDS, SAS PSG-objective sleep measures | EEG-fMRI | 10 |
| Deng et al., 2022 [69] | Between-group comparison; Correlation | 46 HC  23 melancholic MDD-LI 30 melancholic MDD-HI | DSM-IV | MDD-LI: 83% medication  MDD-HI: 63% medication | Depressive episode | HAMD-17, HAMA cutoff for insomnia level:  sleep scores in HAMD >= or < 3 | rsfMRI | 10 |
| Gong et al., 2021a [32] | Correlation | 70 ID  63 good sleepers | ICSD-3 | Not use at least 2 weeks | - | PSQI, SDS, SAS | rsfMRI | 10 |
| Gong et al., 2021b [54] | Correlation | 42 ID  33 HC | ICSD-3 | Not use at least 2 weeks | - | PSQI, SAS, SDS | rsfMRI | 8 |
| Yu et al., 2018 [40] | General linear model | 65 MDD  67 ID | DSM-IV | Naïve to medications;  Or washout period of at least 5.5 lives of medicine | - | HAMD, HAMA, SAS, SDS, PSQI | sMRI | 9 |
| Gong et al., 2023a [60] | MVPA  SVR  (ML) | 60 ID  60 good sleepers | ICSD-3 | Not use at least 2 weeks | - | PSQI, SDS, SAS | rsfMRI | 10 |
| Zhu et al., 2021[47] | Correlation | 27 ID  32 HC | DSM-IV | Not use at least 3 months | - | SAS, SDS | rsfMRI | 10 |
| Xu et al., 2024[56] | Between-group comparison | 57 ID comorbid GAD;  57 HC | ICSD-3; DSM-5 | Not use at 2 weeks | - | PSQI, ISI, SAS, HAMA | sMRI; rsfMRI | 9 |
| Lv et al., 2023[65] | Between-group comparison; SVM (ML) | 60 MDD  34 HC | DSM-5 | - | - | HAMD-17; BAI | rsfMRI | 9 |
| Yin et al., 2024[59] | Correlation | 30 ID  30 HC | DSM-5 | - | - | PSQI, HAMD, HAMA | rsfMRI | 9.5 |
| Shen et al., 2024[55] | Between-group comparison | 51 ID with anxiety; 51 ID without anxiety | ICSD-3 | - | - | PSQI, ISI, SDS, SAS | rsfMRI | 9.5 |
| Xu et al., 2023[51] | Between-group comparison;  Correlation;  Mediation analysis | 44 patients with ID comorbid MDD;  43 HC | DSM-5 | Not use for 2 weeks | - | PSQI, ISI, HAMD, BDI | ASL MRI, rsfMRI | 9 |
| Hu et al., 2024[41] | Between-group comparison;  Correlation | 38 MDD, 37 ID, 49 patients with comorbid depression and insomnia, 50 HC | ICD-10 | x | First episode; | HAMD-17, PSQI, ISI | rsfMRI | 10 |
| Wang et al., 2024[27] | Between-group comparison; Correlation | Dataset1: 56 ID, 53 HC;  Dataset2: 35 ID, 35 HC | Dataset1: ICSD-3;  Dataset2: DSM-5 | No use for 2 weeks | - | PSQI, ISI, BDI | rsfMRI | 10 |
| Lu et al., 2023[66] | Between-group comparison | 55 MDD with insomnia,  39 MDD without insomnia symptoms, 47 HC | DSM-5 | Medication-naïve | - | HAMD-24, HAMA, YMRS, PSQI | H-MRS | 10 |
